# Evaluation of anti-SARS-CoV-2 antibody response to COVID-19 vaccine deployment in the Bono Region, Ghana

**DOI:** 10.1101/2022.06.09.22276192

**Authors:** Samuel Fosu Gyasi, Emmanuel Timmy Donkoh, Samuel Frempong, Akwasi Asamoah, Abdul Sakibu Raji, Rabbi Coffie Baidoo, Williams Isaac, Dorcas Essel, Herbert Alemiya Asakiya

## Abstract

**Background:** Preliminary data across the globe shows that the AstraZeneca vaccine was highly effective in preventing not only the symptoms but also the transmission of the SARS-CoV-2 virus. In Ghana, data on the immune response generated by different vaccination doses is lacking. The present study aimed to compare the anti-SARS-CoV-2 antibody response among single and double-vaccinated versus unvaccinated individuals.

**Methods:** A case-control design was employed for this study. Seventy-nine participants (35 vaccinated, 44 unvaccinated) were recruited from the Sunyani West Municipality and screened for the presence of SARS-CoV-2 specific IgG and IgM antibodies in plasma samples using a Standard COVID IgG and IgM Combo FIA test. Data analysis was carried out with STATA (Version 21).

**Results:** The current study showed that mean IgG levels among vaccine groups (Group-1: Not vaccinated, Group-2: 1 dose, Group-3: 2 doses) differed significantly (F_2, 76_=11.457, p<.001) between Group-1 and Group-3; and between Group-2 and Group-3. Participants in Group-2 and Group-3 were 4.1 and 12.5 times more likely to develop more antibody responses compared to their counterparts in Group-1 respectively.

**Conclusion:** The study showed that participants who took one shot of the vaccine, as well as those who took two shots of the AstraZeneca Vaccine, were 4.1 and 12.5 times more likely to develop a greater antibody response than those who did not receive the vaccine respectively.

## INTRODUCTION

The emergence of COVID-19 caused by an enclosed, single-stranded, positive-sense RNA virus (SARS-CoV-2) in Wuhan, China and its subsequent rise to a global pandemic has had a tremendous impact on people’s lives, causing severe disruption in health systems and having a significant influence on the worldwide economy (1). More than 426,624,859 confirmed cases of COVID-19 have been reported worldwide, with 5,899,578 deaths. So far, Africa has reported 8,303,144 cases with 169,288 deaths and Ghana has had over 159,006 confirmed cases of COVID-19 with 1,442 deaths (WHO, 2021b). The unusual spike in cases worldwide is thought to be due to limited pre-existing immunity(2).

Vaccination has historically proven to be one of the most effective methods of disease burden alleviation and subsequent elimination (3-5). As of January 12, 2022, the WHO had reviewed and approved nine COVID-19 vaccines for emergency use: AstraZeneca/Oxford, Johnson & Johnson, Moderna, Pfizer/BioNTech, Sinopharm, Sinovac, Covaxin, Covovax, and Nuvaxovid (6). Currently, six vaccines have been approved in Ghana: these are [1] AZD1222 Covishield (Oxford/AstraZeneca); [2] ChAdOx1 nCoV-19 Vaxzevria (Oxford/AstraZeneca); [3] JNJ-78436735 Janssen (Johnson & Johnson), [4] Gam-COVID-Vac Sputnik V (Gamaleya); [5] BNT162b2 Comirnaty (Pfizer/BioNTech); [6] mRNA-1273 Spikevax (Moderna), with the AstraZeneca/Oxford vaccine being the most widely deployed (7).

Vaccination provides population-level immunity to an infectious disease by creating an immunological memory for a particular pathogen and inducing the production of protective antibodies. As with other coronaviruses, the spike protein of SARS-CoV-2 is required for viral attachment, fusion, entrance, and transmission. As such they serve as an ideal target for vaccine development aimed at minimizing the spread of the disease (8-11). As a result, the antibody response to spike protein post-vaccination is critical for adjudging the ‘effectiveness’ of the vaccine (12). The detectable levels of IgM and IgG antibodies could provide information regarding seroconversion, as the detection of IgM antibodies indicates a recent exposure whereas the detection of IgG antibodies in the absence of detectable IgM antibodies indicates long-term immunity against the virus (13). Results from COVID-19 AstraZeneca vaccine clinical trials suggest that a single dose elicits an increase in spike-specific antibodies by day 28 and neutralizing antibodies in all participants after a booster dose (14, 15).

There is a question about the effectiveness of the antibody response among vaccinated and unvaccinated individuals in the African setting where the disease has not exacted as much a toll as it has on other continents. However, very few studies have been undertaken to investigate the humoral immune response to SARS-CoV-2 vaccines, specifically the AstraZeneca vaccine widely deployed in Sub-Saharan Africa. In Ghana, studies assessing IgG and IgM antibody responses to the AstraZeneca COVID-19 vaccination are severely lacking. The goal of this study, therefore, was to compare the anti-SARS-CoV-2 antibody response among single and double-vaccinated versus unvaccinated individuals in a peri-urban sub-Saharan community.

## MATERIALS AND METHODS

### Study design and setting

A case-control study of unmatched outcomes was conducted in the Sunyani West Municipality of the Bono region of Ghana from November 2021 to November 2022 among individuals receiving the SARS CoV-2 vaccine (AZD1222, Oxford/AstraZeneca, Pune, India) and control treatments (unvaccinated individuals) with 1 control per experimental subject. The vaccine was administered as a 0.5 ml dose given intramuscularly into the deltoid muscle (upper arm). Data available to our lab from the University community before mass vaccination campaigns in the region indicates that the probability of seroconversion among unvaccinated individuals is 0.2. If the true probability of seroconversion among vaccinated individuals is 0.6 (16), we will need to study 34 case patients and 34 control patients to be able to reject the null hypothesis that the seropositivity rates for vaccinated and unvaccinated controls are equal with probability (power) 0.9. The Type I error probability associated with this test of this null hypothesis is 0.05. A continuity-corrected chi-squared statistic or Fisher’s exact test will be used to evaluate this null hypothesis at baseline and during follow-up.

Community members heard about the research through public announcements on radio and in community fora. Individuals with PCR-confirmed prior infection, pregnant women, individuals younger than 18 years, or seriously ill persons were not included in the study.

### Participants and Data collection

A detailed schematic of the study is provided in Figure 1. Study participants were recruited using a multistage cluster sampling protocol in which the entire municipal health directorate was mapped into clusters/enumeration areas (EA) based on data from the 2021 Population and Housing Census. With support from the Ghana Health Service, three clusters were selected based on vaccination coverage and another three unvaccinated clusters as controls. In each cluster, housing units with eligible individuals were selected as follows: large sparse settlements were sub-divided into four sectors using a sketch map and major landmarks (e.g. road, river, school, mosque, church, etc.). One sector was randomly selected and its centre was identified. From the centre, a first housing unit was identified by pen-spinning. Successive housing units were identified by serpentine movement. Smaller settlements did not require an initial sub-division step.

**Figure 1:**
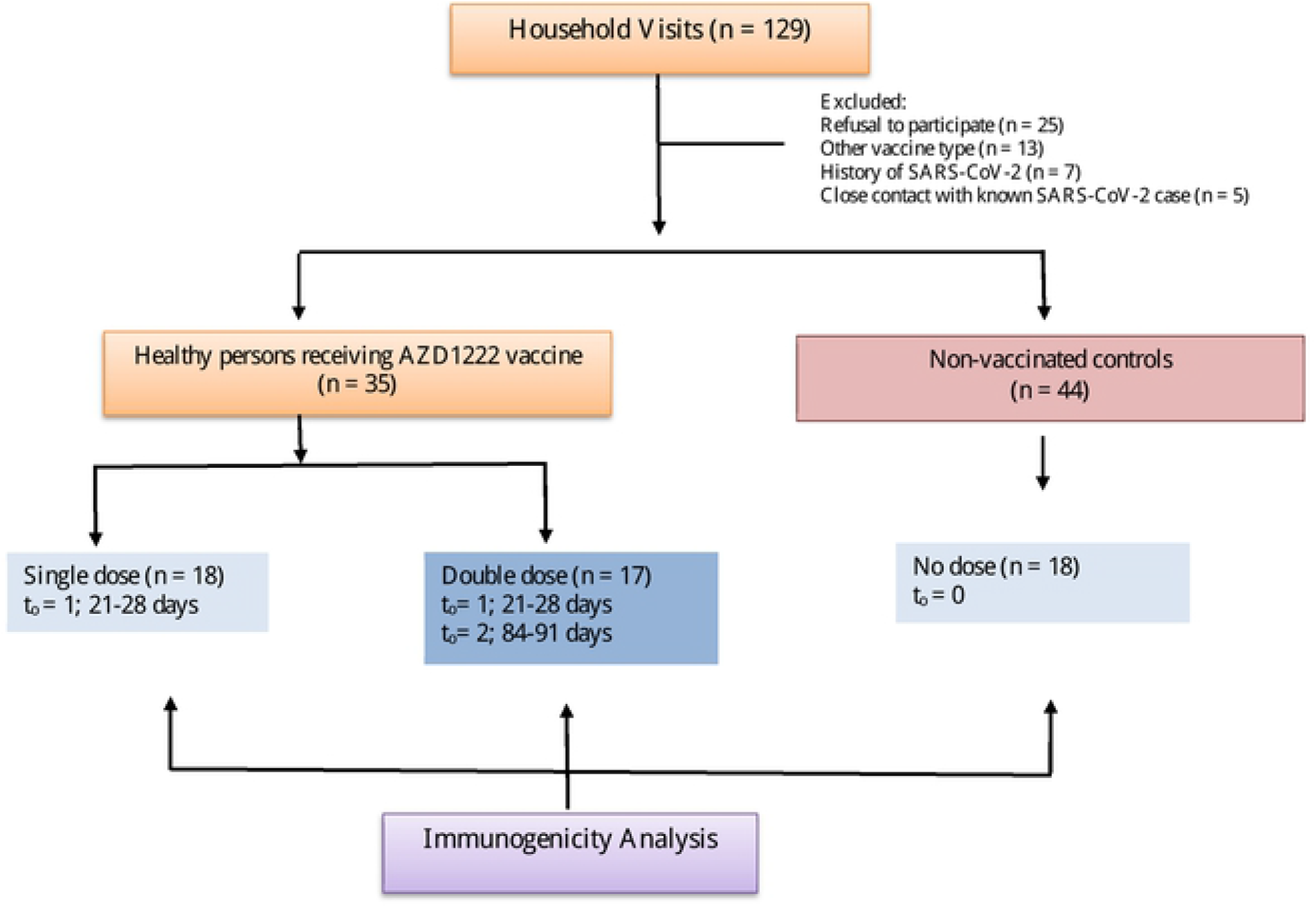
Flow diagram illustrating nested case control design.

The study was explained to all members of each housing unit and random number tables were used to select a single eligible individual per housing unit for documentation of consent. An interviewer-administrated questionnaire (designed using Kobo toolbox data collection tool version 2021.3.4 in English) was used to capture participants’ sociodemographic characteristics. Questions were asked in simple language to ensure proper comprehension before answers were provided.

### Blood Sample Collection and Laboratory Analysis

Venous blood (2-3ml) was collected via venipuncture into labelled EDTA tubes from eligible participants by an experienced phlebotomist at two prespecified time points: from day 21 to 28 (t_o_= 0) and from day 84 to 91 (t_o_=1). The samples collected from participants were transported on ice to the Centre for Research in Applied Biology (CeRAB), centrifuged at 4000rmp for 5mins to separate the plasma into pre-labelled cryotubes and stored at -20°C. Laboratory detection of anti-SARS-CoV-2 S1 spike protein-specific IgG and IgM antibodies was performed using the STANDARD F COVID-19 IgM / IgG Combo FIA test kits on the F2400 Analyzer (SD Biosensor, Inc., KOREA) according to the manufacturer’s guidelines.

The STANDARD F COVID-19 IgM / IgG Combo FIA detects IgG and IgM antibodies developed against the SARS-CoV-2 S1 spike protein and has a sensitivity of 94.41% and specificity of 90.62% (17). A quantitative measure of the fluorescent signal emitted corresponding to the amount of antibody present provides a basis for establishing seropositivity after establishing a threshold/cut-off index. A cut-off index (COI) equal to or greater than 1.0 indicates a positive result for anti-S1 SARS-CoV-2 IgG and IgM. The antibody index is a numerical representation of the measured fluorescence signal given off by the test reaction that took place within the F2400 Analyzer.

### Ethical Consideration

All research activities and protocols were approved by the Committee for Human Research and Ethics at the University of Energy and Natural Resources with reference number CHRE/AP/09/021. The purpose of the study and all procedures were explained to participants before consent was taken. All participants were provided with informed consent forms and were included after the endorsement of the form.

### Data Analysis and Processing

Data was entered into Microsoft Excel 2016 for initial processing and analyzed with STATA (Version 26). Mean and Standard deviation for quantitative variables were calculated along with frequencies and percentages for all categorical variables. Multiple-way ANOVA tests were performed to estimate the difference in antibody levels between individuals vaccinated with one dose, two doses and unvaccinated individuals. The association between variables such as sex, vaccination status, anti-S1 IgM, and IgG response were evaluated with the Chi-square test of independence. Where required, anti-S1 IgG and IgM indices were compared across categories by the Student’s t-test. Logistic regression was performed to determine the odds of anti-S1 IgG seropositivity across different dose categories. A p-value less than 0.05 was considered statistically significant.

## RESULTS

### Socio-Demographic information on participants

At baseline 35 vaccinated and 44 non-vaccinated individuals fulfilled all criteria and were eligible for follow-up. Baseline characteristics of the study population are shown in Table 1. The mean age was 32 ±14.3 years. Participants were mostly males (65.8%). Common complaints post-vaccination included pain at injection site (62.9%), headache (37.1) and malaise (25.8).

**Table 1:** Baseline Characteristics of Study Participants.

| Variable | n (%) |
| --- | --- |
| *Mean Age (SD)* | 32 ± 14.3 |
| *Sex (n=79)* |  |
| Males | 52 (65.8) |
| Females | 27 (34.2) |
| *Vaccination Status (n=79)* |  |
| None | 44 (55.7) |
| One dose | 18 (22.8) |
| Two doses | 17 (21.5) |
| *Post-vaccination adverse events (N=35)* |  |
| Yes | 25 (71.4) |
| No | 10 (28.6) |
| *Common complaints (N=35)* |  |
| Pain at injection site | 22 (62.9) |
| Headache | 13 (37.1) |
| Fever | 8 (22.9) |
| General malaise | 9 (25.8) |
| Chills | 10 (28.6) |
| Arthralgia | 1 (2.9) |

Footnote: N= 79

### Prevalence of anti-S1 IgM and IgG among participants

Baseline data on seroconversion to anti-S1 protein IgM and IgG for vaccinated and individuals and controls are presented in Table 2. Among non-vaccinated individuals (13/44; 29.5%) anti-S1 IgM seropositivity was higher than was recorded for single (4/18; 22.2%) and double vaccinated (1/17; 5.9%) individuals. Anti-S1 IgG titres were much higher across all three vaccination categories. However, non-vaccinated individuals (24/44; 54.5%) had significantly lower anti-S1 IgG titres compared to single (15/18; 83.3%) and double vaccinated individuals (16/17; 94.1%) [*x*^2^ (df=2, *N*= 79) = 11.15, *p* = .004].

**Table 2:** Prevalence of IgM and IgG anti-S1 antibody among participants.

| Parameter | Anti-S1 IgM |  | X<sup>2</sup>-<br>Value, df | P-Value | Anti S1 IgG |  | X<sup>2</sup>-<br>Value, df | P-Value |
| --- | --- | --- | --- | --- | --- | --- | --- | --- |
|  | Positive | Negative |  |  | Positive | Negative |  |  |
| Vaccination Status |  |  |  |  |  |  |  |  |
| No dose | 13 (29.5) | 31 (70.5) | 3.91, 2 | 0.142 | 24 (54.5) | 20 (45.5) | 11.15, 2 | 0.004 |
| Single dose | 4 (22.2) | 14 (77.8) |  |  | 15 (83.3) | 3 (16.7) |  |  |
| Double dose | 1 (5.9) | 16 (94.1) |  |  | 16 (94.1) | 1 (5.9) |  |  |
| Total | 18 (22.8) | 61 (77.8) |  |  | 55 (69.6) | 24 (30.4) |  |  |

Note: Anti-S1 IgM: Anti SARS-CoV-2 spike protein 1 immunoglobulin M, Anti-S1 IgG: Anti SARS-CoV-2 spike protein 1 immunoglobulin G, X<sup>2</sup>: Chi-square value, df: Degree of freedom. P-value considered significant at p<0.05.

To test the null hypothesis that antibody responses are similar across the various levels of vaccine dose, participants were divided into 3 (group 1: Not vaccinated, Group 2: 1 dose, Group 3: 2 doses) and mean titres compared by ANOVA (Table 3). Significant differences in the levels of antibody response were observed (F_2, 76_=11.46, p<.001). Post-hoc comparisons to reveal heterogeneity between groups were performed using Dunnett’s T3 as shown in Table 3 below. The mean COI for the unvaccinated group (M=6.19, SD=6.42) was significantly lower compared to the vaccinated group. However, the single-dose group’s (M=10.75, SD=6.39) antibody response did not differ significantly from the double-dose group (M=13.88, SD=3.56).

**Table 3:** Number of Doses Stratified by Test of Homogeneity of Variance.

| *No. of doses* | **Test of Homogeneity of Variance** |  | **ANOVA** |  |
| --- | --- | --- | --- | --- |
|  | *Mean* | *SD* | *F* | *Sig.* |
| None | 6.19 | 6.42 | 11.46 | 0.000 |
| 1st Dose | 10.75 | 6.39 |  |  |
| 2nd Dose | 13.87 | 3.56 |  |  |

| **Differences between groups** |  |  |  |
| --- | --- | --- | --- |
| *No. of doses* | *Mean Difference* | *Sig.* | *95% Confidence interval* |
| None- 1 dose | -4.56 | 0.046 | -9.06. - -0.06 |
| None- 2 doses | -7.68 | 0.000 | -10.88 - -4.48 |

Logistic regression was performed to determine the odds of anti-S1 IgG seropositivity across different dose categories (Table 4). Double vaccinated individuals were 4.1 times more likely to develop anti-S1 IgG seropositivity compared to unvaccinated individuals. Again, those who took two doses were 12.5 times more likely to develop anti-S1 IgG seropositivity than those without the vaccine.

**Table 4:** Odds of anti-S1 IgG seropositivity.

| **Antibodies level** | **AOR (95%)** | **P-Value** | **CI** |
| --- | --- | --- | --- |
| **No. of Doses** |  |  |  |
| None | 1 | - | - |
| One dose | 4.10 | **0.044\*** | 1.04 - 16.27 |
| Two doses | 12.50 | **0.021\*** | 1.48 - 105.52 |

*Footnote: \* p-value<0.05, AOR: Adjusted Odds Ratio, CI: Confidence Interval,*

## DISCUSSION

To the best of our knowledge, this is the first report from Ghana to report data on the SARS-CoV-2 antibody response following widespread community administration of Covishield by the Ghana Health Service. Community vaccination does not only provide individual protection, but also facilitates the attainment of herd immunity to protect immunocompromised, unvaccinated and immunologically naïve individuals by lowering the number of vulnerable people below the transmission threshold (18). Despite the high efficacy of most currently approved vaccines, particularly in reducing the risk of clinical aggravation (19) accumulating data show significant heterogeneity in the immune response elicited after vaccination among different individuals (18).

The acute phase antibody (IgM) showed no significant association with vaccination status (Table 2). A reason for this observation is that the anti-SARS-CoV-2 S1 IgM antibody can be acquired through either natural infection or vaccination and is expected to wane rapidly over time (20, 21). Therefore, in areas where viral exposure and vaccine coverage are high, anti S1 IgM antibody titres may not vary significantly among these groups.

On the other hand, seroconversion to the chronic phase antibody (IgG) was associated with significant variation among vaccination categories as reported in previous vaccine efficacy studies (22). IgG levels are known to be critical in ensuring protection against viral diseases (23). Also, IgG antibodies have a longer half-life and are the most commonly used in clinical settings as a marker of long-term protection or vaccine efficacy (24).

Significant differences in the levels of antibody response were observed (Table 3). Post-hoc comparisons revealed that antibody titres for the unvaccinated group were significantly low when compared to the two vaccinated groups. However, we did not observe significant differences in antibody titre between single vaccinated and double vaccinated individuals at baseline. This phenomenon could be attributed to the time of sampling being too premature in the time course of vaccination for any variability to be observed and can be clarified during follow-up (25). However, in regression analysis, single and double vaccinated individuals were respectively 4.1 and 12.5 times more likely to have positive IgG antibody titres than vaccine-naïve participants (Table 4).

## CONCLUSION

We have assessed the IgM and IgG SARS-CoV-2 spike protein antibody response between vaccinated (single and double dose) and unvaccinated individuals in peri-urban communities in Ghana. Initial results demonstrate that in the short-term taking one or two doses of the AstraZeneca vaccine generates a significant protective antibody response over not taking the vaccine at all. It remains to be seen how long the generated immune response lasts will last in this population and whether a booster shot could be a useful strategy.

## DECLARATIONS

### CONSENT FOR PUBLICATION

All authors have duly consented for this manuscript to be submitted for publication

### AVAILABILITY OF DATA AND MATERIALS

The data for this study is available in the manuscript and in its attached Supplementary File: **S1 File. Study Data**. It can also be obtained through the Committee for Human Research and Ethics (CHRE) of the University of Energy and Natural Resources.

### AUTHORS’ CONTRIBUTION

SFG conceived and designed the study and provided supervision, ETD contributed to the study design, provided supervision and drafted the manuscript with AA. AA, ASR and SF undertook sample collection, processing and performed laboratory analysis. RCB, DE and WI undertook participant recruitment, field data collection and assisted with experimental testing in the laboratory. HAA analyzed the data and reported the findings. All authors reviewed and approved the final manuscript.

## ACKNOWLEDGEMENTS

We wish to acknowledge the chiefs and people of the Fiapre community for their support during this work.

## REFERENCES

1. Singh S, Kumar R, Panchal R, Tiwari MK. Impact of COVID-19 on logistics systems and disruptions in food supply chain. International Journal of Production Research. 2021;59(7):1993–2008.

2. Sette A, Crotty S. Author Correction: Pre-existing immunity to SARS-CoV-2: the knowns and unknowns. Nature reviews Immunology. 2020;20(10):644.

3. Bloom DE. The value of vaccination. Hot topics in infection and immunity in children VII: Springer; 2011. p. 1–8.

4. Echeverria-Londono S, Li X, Toor J, de Villiers MJ, Nayagam S, Hallett TB, et al. How can the public health impact of vaccination be estimated? BMC Public Health. 2021;21(1):1–12.

5. Preaud E, Durand L, Macabeo B, Farkas N, Sloesen B, Palache A, et al. Annual public health and economic benefits of seasonal influenza vaccination: a European estimate. BMC Public Health. 2014;14:813.

6. WHO. COVID-19 advice for the public: Getting vaccinated 2022 [Available from: https://www.who.int/emergencies/diseases/novel-coronavirus-2019/covid-19-vaccines/advice.

7. Baffoe GA, Nortey NKLW. An Analysis of Conspiracy Beliefs of Covid-19 Vaccination in Ghana. Advances in Journalism Communication. 2021;9(4):196–208.

8. Du L, He Y, Zhou Y, Liu S, Zheng B-J, Jiang S. The spike protein of SARS-CoV — a target for vaccine and therapeutic development. Nature Reviews Microbiology. 2009;7(3):226–36.

9. Du L, Yang Y, Zhou Y, Lu L, Li F, Jiang S. MERS-CoV spike protein: a key target for antivirals. Expert opinion on therapeutic targets. 2017;21(2):131–43.

10. Wu W, Wang A, Liu M. Clinical features of patients infected with 2019 novel coronavirus in Wuhan, China. Lancet. 2020;395(10223):497–506.

11. Jiang S, Hillyer C, Du L. Neutralizing Antibodies against SARS-CoV-2 and Other Human Coronaviruses. Trends in immunology. 2020;41(5):355–9.

12. Raturi M, Kusum A, Kala M, Mittal G, Sharma A, Bansal N. Locally harvested Covid-19 convalescent plasma could probably help combat the geographically determined SARS-CoV-2 viral variants. Transfusion clinique et biologique : journal de la Societe francaise de transfusion sanguine. 2021;28(3):300–2.

13. Jamiruddin R, Haq A, Khondoker MU, Ali T, Ahmed F, Khandker SS, et al. Antibody response to the first dose of AZD1222 vaccine in COVID-19 convalescent and uninfected individuals in Bangladesh. Expert review of vaccines. 2021;20(12):1651–60.

14. Falsey AR, Sobieszczyk ME, Hirsch I, Sproule S, Robb ML, Corey L, et al. Phase 3 Safety and Efficacy of AZD1222 (ChAdOx1 nCoV-19) Covid-19 Vaccine. The New England journal of medicine. 2021;385(25):2348–60.

15. Folegatti PM, Ewer KJ, Aley PK, Angus B, Becker S, Belij-Rammerstorfer S, et al. Safety and immunogenicity of the ChAdOx1 nCoV-19 vaccine against SARS-CoV-2: a preliminary report of a phase 1/2, single-blind, randomised controlled trial. The Lancet. 2020;396(10249):467–78.

16. Suhandynata RT, Hoffman MA, Kelner MJ, McLawhon RW, Reed SL, Fitzgerald RL. Longitudinal monitoring of SARS-CoV-2 IgM and IgG seropositivity to detect COVID-19. The journal of applied laboratory medicine. 2020;5(5):908–20.

17. Biosensor S. Standard F COVID-19 IgM/IgG Combo FIA. In: Biosensor S, editor. Package insert2019.

18. Mallory ML, Lindesmith LC, Baric RS. Vaccination-induced herd immunity: successes and challenges. Journal of Allergy Clinical Immunology. 2018;142(1):64–6.

19. Doroftei B, Ciobica A, Ilie O-D, Maftei R, Ilea C. Mini-review discussing the reliability and efficiency of COVID-19 vaccines. Diagnostics. 2021;11(4):579.

20. Jalkanen P, Kolehmainen P, Häkkinen HK, Huttunen M, Tähtinen PA, Lundberg R, et al. COVID-19 mRNA vaccine induced antibody responses against three SARS-CoV-2 variants. Nature communications. 2021;12(1):1–11.

21. Banga Ndzouboukou JL, Zhang YD, Lei Q, Lin XS, Yao ZJ, Fu H, et al. Human IgM and IgG Responses to an Inactivated SARS-CoV-2 Vaccine. Curr Med Sci. 2021;41(6):1081–6.

22. Wei J, Stoesser N, Matthews PC, Ayoubkhani D, Studley R, Bell I, et al. Antibody responses to SARS-CoV-2 vaccines in 45,965 adults from the general population of the United Kingdom. Nat Microbiol. 2021;6(9):1140–9.

23. Murin CD, Wilson IA, Ward AB. Antibody responses to viral infections: a structural perspective across three different enveloped viruses. Nature microbiology. 2019;4(5):734–47.

24. Racine R, Winslow GM. IgM in microbial infections: taken for granted? Immunology letters. 2009;125(2):79–85.

25. Hou H, Wang T, Zhang B, Luo Y, Mao L, Wang F, et al. Detection of IgM and IgG antibodies in patients with coronavirus disease 2019. Clinical translational immunology. 2020;9(5):e1136.

